# Inflammation and postoperative complications after major surgery: a bi-directional two-sample Mendelian randomization study

**DOI:** 10.64898/2026.03.26.26349385

**Authors:** Richard A. Armstrong, Paul Yousefi, Ben Gibbison, Golam M Khandaker, Tom R Gaunt

## Abstract

**Background:** Observational studies have reported an association between inflammation and postoperative complications but it is unclear whether these associations are causal. It is also unknown whether postoperative outcomes share a causal architecture with chronic, all-cause disease.

**Methods:** We performed bi-directional two-sample Mendelian randomization to investigate potential causal effects of 19 genetically-proxied inflammatory markers on postoperative acute kidney injury, atrial fibrillation (AF), delirium, myocardial infarction, stroke and surgical site infection, and their all-cause equivalents. Genetic instruments for inflammatory markers were sourced from nine GWAS of up to 204,402 European participants with outcome data derived from UK Biobank.

**Results:** The primary postoperative analysis showed a protective effect of down-regulated IL-6 signalling on stroke risk (OR (95% CI) 0.27 (0.11–0.69), p=0.006). However, in the all-cause analysis a causal effect on stroke was not present (OR (95% CI) 1.14 (0.75–1.24), p=0.78), whilst a robust protective effect was seen for down-regulated IL-6 with AF across all three instruments studied (all p<0.009). In postoperative and all-cause analyses, genome-wide variants for CRP showed a protective effect on delirium that was not present in *cis*-restricted analyses.

**Conclusions:** We found evidence supporting a potential causal role for IL-6 signalling in perioperative stroke. However, the divergence in IL-6 effects between postoperative and all-cause outcomes suggests that the inflammatory architecture of acute postoperative complications may differ from chronic disease states. Furthermore, our findings suggest previously reported associations between CRP and delirium likely represent horizontal pleiotropy rather than direct causation. Future work should interrogate local tissue responses and the immediate perioperative period.

## Introduction

Postoperative complications after major surgery occur in up to 15% of patients worldwide with substantial impacts on morbidity, mortality and resource utilisation ^1^. Major surgery produces an inflammatory response as part of normal adaptive and healing processes, but this can become dysregulated, resulting in varied clinical phenotypes depending on the organ system affected. Observational associations have been reported between systemic inflammation, as proxied by a broad range of inflammatory markers, and complications including acute kidney injury (AKI) ^2, 3^, atrial fibrillation (AF) ^4, 5^, myocardial infarction (MI) ^6^, delirium ^7^, surgical site infection (SSI) ^8^ and stroke ^9^. A causal relationship is biologically plausible, supported by mechanistic theories including the neuroinflammatory hypothesis of delirium ^10^, the direct effect of inflammatory mediators on cardiac conductive tissue in AF ^11^, and the pro-thrombotic state associated with inflammation in the context of MI and stroke ^6, 12^.

However, most existing studies cannot distinguish correlation from causation and are not designed to test evidence of causality between inflammation and postoperative complications. If inflammation is indeed causal, this may suggest potential treatment strategies, for example immunomodulatory therapies. Mendelian randomization (MR) is one approach to address the causal hypothesis by using genetic variants as proxies for the exposure of interest. Compared to other observational methods, MR is less susceptible to unmeasured confounding and reverse causation as genetic variants are randomly assigned and fixed at conception ^13^.

Previous MR studies have examined the role of inflammatory markers in nonpostoperative phenotypes related to the postoperative complications above. For example, there is evidence supporting a causal role for IL-6 in coronary artery disease (CAD) ^14, 15^, stroke ^15, 16^ and AF ^15^ and for MCP-1 in CAD ^17^ and stroke ^18^, whilst CRP has shown null effects in CAD ^19^, stroke ^20^, AF ^21^, renal function ^22^ and delirium ^23^. It is unclear, however, whether the postoperative occurrence of these phenotypes, in the context of the acute systemic inflammatory response to major surgery, involves the same causal pathways as their chronic development over the life course.

We undertook a two-sample MR study to systematically evaluate possible causal effects of genetically predicted circulating levels of inflammatory markers on the risk of six major postoperative complications: AF, AKI, MI, delirium, stroke and SSI. Immunological proteins were selected based on their reported associations with these complications in previous observational studies^2–8, 10, 11, 24–33^. The MR approach allows an interrogation of whether these proteins represent upstream causal pathways, and hence potential therapeutic targets, or are simply acting as downstream biomarkers of a disease process. Bi-directional MR was conducted, with the inflammatory markers as outcomes, to test the direction of association. We then extended our analyses to non-postoperative outcomes to explore potential differences between the acute and chronic onset of these conditions.

## Methods

### Exposures: genetic variants related to inflammatory markers

Mendelian randomization (MR) uses genetic variants associated with exposures and outcomes as proxies. For an instrument to be valid it must be: 1) reliably associated with the exposure (relevance assumption); 2) associated with the outcome only through the exposure (independence assumption); 3) independent of unobserved confounders that influence both exposure and outcome (exclusion-restriction) ^34^.

The inflammatory markers included in the study were selected due to reported association with at least one of the postoperative complications of interest (Table 1, Supplementary Table 1). Genetic variants were identified from summary statistics of large GWAS of European participants. All variants associated with the outcome trait at a p value threshold of p < 10^−5^ and independent of each other with a clumping window of 10,000 kb and r^2^ threshold of < 0.001 were extracted as genome-wide/*trans* variants. Where present, *cis* variants were defined as those within a ±1Mb window of the protein coding gene region based on Genome Reference Consortium Human Build 37. *Cis*-variants were considered the primary instruments for testing specific causal hypotheses for individual biomarkers. These variants are biologically anchored to the specific protein coding region and so are less likely to have pleiotropic effects on the outcome of interest via other pathways (Supplementary Methods 1). *Trans*-variants were included as supportive evidence to characterise the broader genetic architecture of the inflammatory response, acknowledging that these may act through indirect pathways (horizontal pleiotropy). Where a SNP was not present in the outcome dataset, proxy SNPs were identified using linkage disequilibrium (LD) tagging (r^2^ > 0.8) (*LDlinkR* ^35^).

**Table 1.**
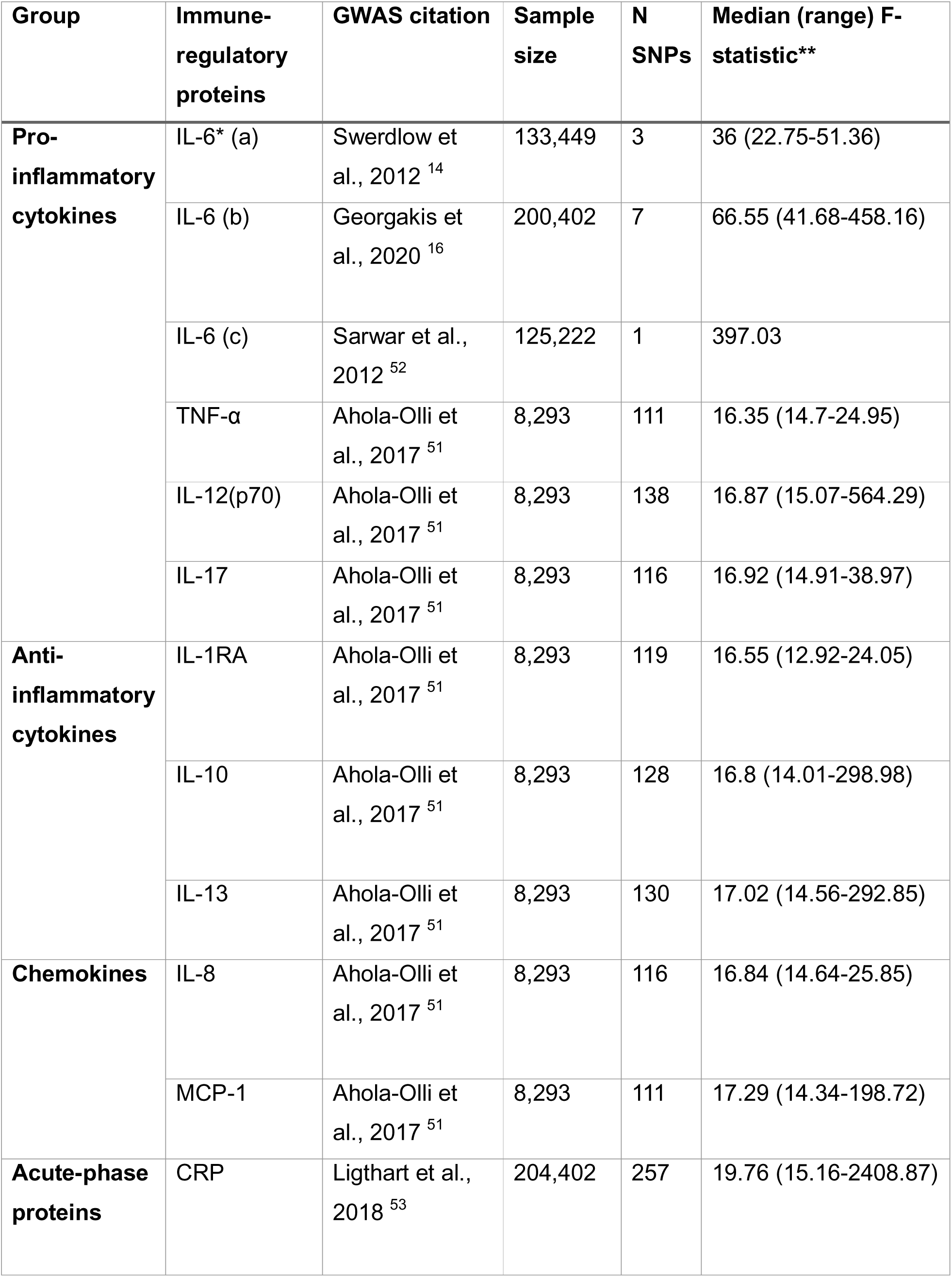
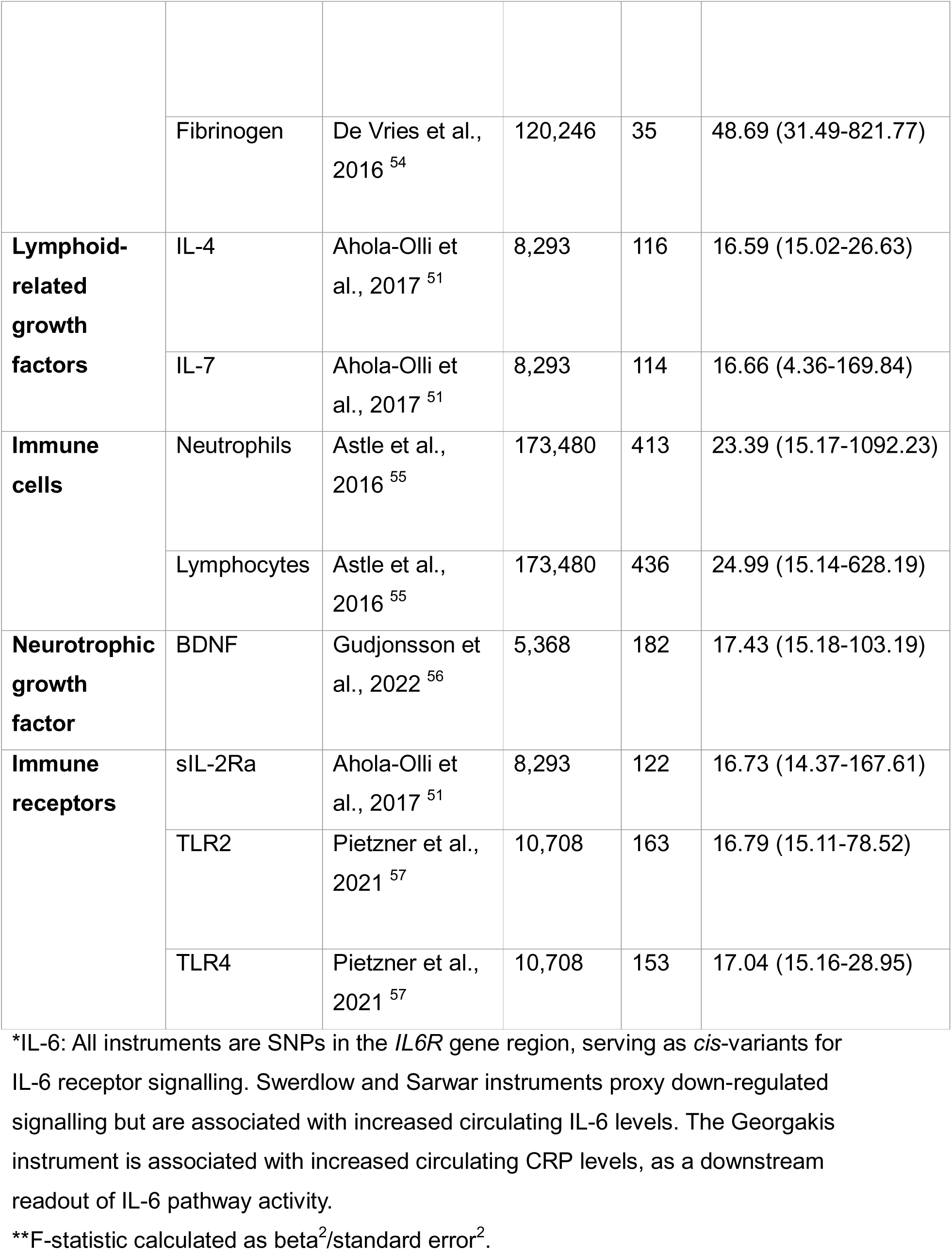
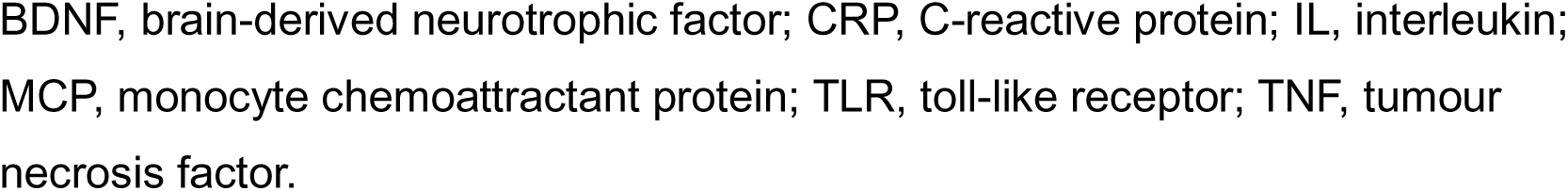
Inflammatory markers included in analyses with citation for GWAS summary statistics.

| **Group** | **Immune-regulatory proteins** | **GWAS citation** | **Sample size** | **N SNPs** | **Median (range) F-statistic\*\*** |
| --- | --- | --- | --- | --- | --- |
| **Pro-inflammatory cytokines** | IL-6* (a) | Swerdlow et al., 2012 <sup>14</sup> | 133,449 | 3 | 36 (22.75-51.36) |
|  | IL-6 (b) | Georgakis et al., 2020 <sup>16</sup> | 200,402 | 7 | 66.55 (41.68-458.16) |
|  | IL-6 (c) | Sarwar et al., 2012 <sup>52</sup> | 125,222 | 1 | 397.03 |
| | TNF-α | Ahola-Olli et al., 2017 <sup>51</sup> | 8,293 | 111 | 16.35 (14.7-24.95) |
|  | IL-12(p70) | Ahola-Olli et al., 2017 <sup>51</sup> | 8,293 | 138 | 16.87 (15.07-564.29) |
|  | IL-17 | Ahola-Olli et al., 2017 <sup>51</sup> | 8,293 | 116 | 16.92 (14.91-38.97) |
| **Anti-inflammatory cytokines** | IL-1RA | Ahola-Olli et al., 2017 <sup>51</sup> | 8,293 | 119 | 16.55 (12.92-24.05) |
|  | IL-10 | Ahola-Olli et al., 2017 <sup>51</sup> | 8,293 | 128 | 16.8 (14.01-298.98) |
|  | IL-13 | Ahola-Olli et al., 2017 <sup>51</sup> | 8,293 | 130 | 17.02 (14.56-292.85) |
| **Chemokines** | IL-8 | Ahola-Olli et al., 2017 <sup>51</sup> | 8,293 | 116 | 16.84 (14.64-25.85) |
|  | MCP-1 | Ahola-Olli et al., 2017 <sup>51</sup> | 8,293 | 111 | 17.29 (14.34-198.72) |
| **Acute-phase proteins** | CRP | Ligthart et al., 2018 <sup>53</sup> | 204,402 | 257 | 19.76 (15.16-2408.87) |
|  | Fibrinogen | De Vries et al., 2016 <sup>54</sup> | 120,246 | 35 | 48.69 (31.49-821.77) |
| **Lymphoid-related growth factors** | IL-4 | Ahola-Olli et al., 2017 <sup>51</sup> | 8,293 | 116 | 16.59 (15.02-26.63) |
|  | IL-7 | Ahola-Olli et al., 2017 <sup>51</sup> | 8,293 | 114 | 16.66 (4.36-169.84) |
| **Immune cells** | Neutrophils | Astle et al., 2016 <sup>55</sup> | 173,480 | 413 | 23.39 (15.17-1092.23) |
|  | Lymphocytes | Astle et al., 2016 <sup>55</sup> | 173,480 | 436 | 24.99 (15.14-628.19) |
| **Neurotrophic growth factor** | BDNF | Gudjonsson et al., 2022 <sup>56</sup> | 5,368 | 182 | 17.43 (15.18-103.19) |
| **Immune receptors** | sIL-2Ra | Ahola-Olli et al., 2017 <sup>51</sup> | 8,293 | 122 | 16.73 (14.37-167.61) |
|  | TLR2 | Pietzner et al., 2021 <sup>57</sup> | 10,708 | 163 | 16.79 (15.11-78.52) |
|  | TLR4 | Pietzner et al., 2021 <sup>57</sup> | 10,708 | 153 | 17.04 (15.16-28.95) |

\*IL-6: All instruments are SNPs in the *IL6R* gene region, serving as *cis*-variants for IL-6 receptor signalling. Swerdlow and Sarwar instruments proxy down-regulated signalling but are associated with increased circulating IL-6 levels. The Georgakis instrument is associated with increased circulating CRP levels, as a downstream readout of IL-6 pathway activity.
\*\*F-statistic calculated as $\beta^2/\text{standard error}^2$ .
BDNF, brain-derived neurotrophic factor; CRP, C-reactive protein; IL, interleukin; MCP, monocyte chemoattractant protein; TLR, toll-like receptor; TNF, tumour necrosis factor.

### Primary outcomes: genetic variants related to postoperative complications

We used summary statistics from case-control GWAS of postoperative AF, AKI, MI, delirium, SSI and stroke performed in UK Biobank ^36, 37^. Participants were included if they underwent major surgery and had a first diagnosis of an ICD-10 coded complication of interest within 30 days of the procedure. Controls were those undergoing major surgery but not experiencing the complication. Individuals were excluded if they had a previous diagnosis of the outcome of interest prior to surgery to reduce potential bias through confounding and reverse causation. This also ensured all events occurred after baseline variables were measured at UK Biobank enrolment and contributed to a more homogenous case-control cohort. To further distinguish between coding of comorbidities and acute events, we used ICD-10 codes specific to acute diagnoses where possible (see Supplementary Methods 2 for further information on inclusion/exclusion criteria and case definition). The numbers of cases and controls for each outcome phenotype are shown in Table 2. Exposure and outcome GWAS were from different consortia, minimising the risk of bias from sample overlap.

**Table 2.** Counts of cases and controls in GWAS of postoperative complications of interest.

| **Complication** | **ICD-10**<br>**diagnosis code** | **Cases** | **Controls** |
| --- | --- | --- | --- |
| Atrial fibrillation | I48.9 | 2,739 | 135,878 |
| Acute kidney injury | N17 | 3,002 | 134,941 |
| Acute myocardial infarction | I21 | 282 | 123,339 |
| Delirium | F05 | 1,016 | 139,148 |
| Stroke | I61, I63, I64 | 569 | 137,842 |
| Surgical site infection | T81.4 | 2,701 | 136,120 |

### Secondary outcomes: genetic variants related to non-postoperative diagnoses

To explore the relationship between postoperative complications and non-postoperative equivalent phenotypes (e.g. postoperative AF and chronic AF) we undertook a parallel analysis including all cases of each ICD-10 phenotype in the UK Biobank cohort (case-control numbers can be found in Supplementary Methods 3). For surgical site infection, an alternative diagnosis of ‘infections of the skin and subcutaneous tissue’ (ICD-10 codes L00-L08) was used. Case-control GWAS adjusted for age, sex, genetic chip and the first ten ancestry principal components was performed for each outcome using regenie (v4.1 ^38^). The UK Biobank study was approved by the North-West Multi-centre Research Ethics Committee and all participants provided written informed consent. This research has been conducted under UK Biobank project number 128619.

### Statistical analysis

Bidirectional two-sample MR was performed using the *TwoSampleMR* package (v. 0.6.25 ^39, 40^) in R (4.5.0). For each instrument, exposure and outcome data are harmonised and palindromic SNPs excluded if strand could not be inferred from MAF (>0.42). Our primary analysis was inverse-variance weighted (IVW) regression if >1 SNP was available or Wald ratio if the instrument comprised only 1 SNP. Covariates such as comorbidities were not adjusted for due to the risk of introducing collider bias within an MR framework (Supplementary Methods 1). Sensitivity analyses included MR-Egger, weighted median, weighted mode and MR Pleiotropy RESidual Sum and Outlier (MR-PRESSO, where outliers were identified). Where these methods required a minimum number of SNPs, they are reported when sufficient SNPs were available. Additionally, Steiger filtering, heterogeneity (Cochran’s Q-statistic) and pleiotropy (Egger intercept) were assessed. Statistical significance thresholds were adjusted for multiple testing using the Bonferroni method according to the number of exposures for each outcome (only those inflammatory markers reported to be associated with each outcome were analysed, Supplementary Table 1).

Code to support the analyses is available on GitHub ^41^. The study is reported as per the Strengthening the Reporting of Observational Studies in Epidemiology using Mendelian Randomization (STROBE-MR) guidelines (see Supplementary data).

## Results

### Inflammation and postoperative outcomes: results from primary analyses

In the primary analysis using *cis*-acting genetic instruments, only genetically proxied down-regulation of IL-6 signalling (via *cis*-variants in the *IL6R* gene region) showed robust evidence of potential causal effect on any postoperative complications. For all other inflammatory markers, we did not find strong evidence of causal effects on the complications of interest after multiple testing correction and sensitivity analyses (Figure 1, Supplementary Table 2).

**Figure 1.**
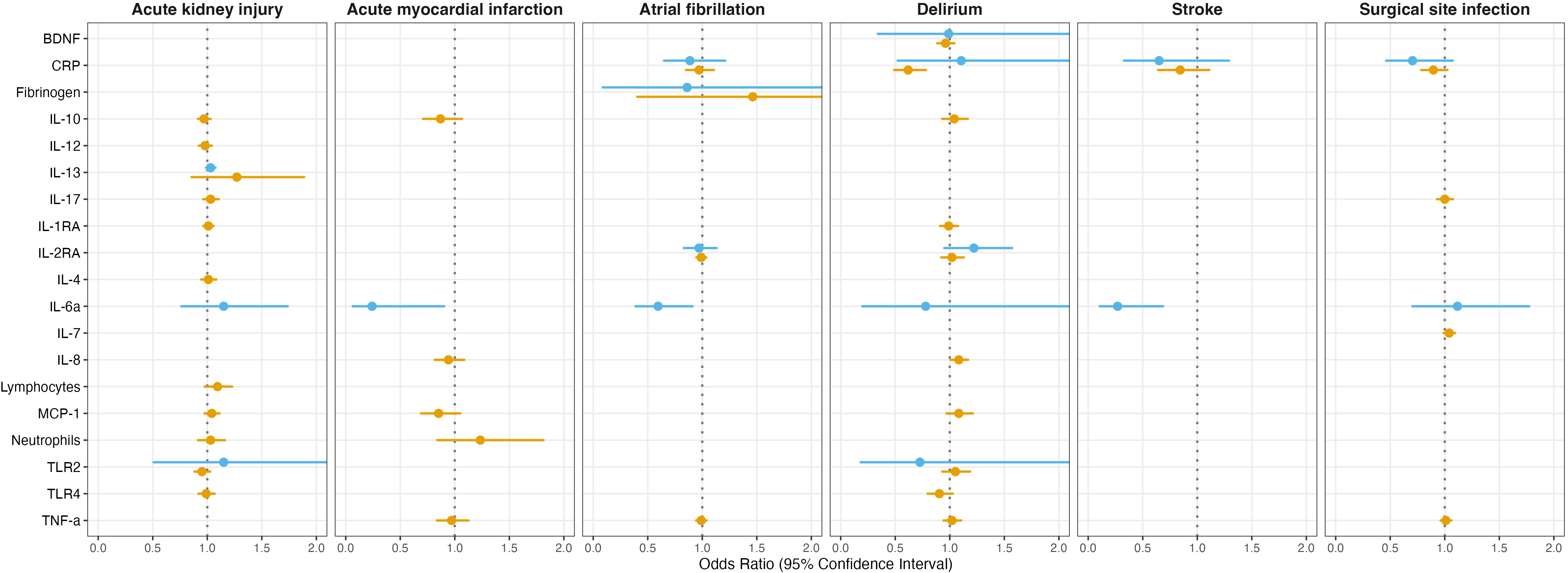
Odds ratios with 95% CIs from Mendelian randomization analysis showing potential causal effects of genetically predicted levels of inflammatory markers on the risks of postoperative complications. Orange dots = trans variants, blue dots = cis variants. BDNF, brain-derived neurotrophic factor; CRP, C-reactive protein; IL, interleukin; IL6a, Swerdlow et al instrument; MCP, monocyte chemoattractant protein; TLR, toll-like receptor; TNF, tumour necrosis factor. X-axes are capped at 2.0; where the upper 95% CI limit was beyond this point the line extends to the edge of the panel.

The Swerdlow et al. instrument for IL-6, which serves as a *cis*-instrument for IL-6 receptor blockade representing reduced IL-6 signalling, demonstrated a protective effect on postoperative stroke risk (IVW OR (95% CI) 0.27 (0.11–0.69), p = 0.006). This finding was consistent across sensitivity analyses, with no evidence of heterogeneity (Cochran’s Q = 0.48, p = 0.79) or directional pleiotropy (MR-Egger intercept (SE) = −0.08 (0.27), p = 0.82), suggesting the causal pathway was indeed via IL-6 receptor blockade rather than an alternative route. Furthermore, there was suggestive evidence (p < 0.05) of the same *cis*-acting IL-6 instrument having a protective effect on the risk of postoperative AF (OR (95% CI) 0.6 (0.39-0.91), p = 0.017) and MI (OR (95% CI) 0.24 (0.06-0.91), p = 0.035). Both suggestive findings were similarly robust to sensitivity analyses, showing no evidence of heterogeneity (Cochran’s Q = 0.48, p=0.79 and 0.01, p=0.99 respectively) or horizontal pleiotropy (MR-Egger intercept (SE) −0.07 (0.12), p = 0.69 and 0.02 (0.39), p = 0.97 respectively).

In the broader genome-wide (*cis* and *trans*) analyses, CRP initially showed a protective effect on postoperative delirium (IVW OR (95% CI) 0.62 (0.49–0.78), p < 0.001). However, sensitivity analyses revealed substantial heterogeneity among the included variants (Cochran’s Q = 348.14, p < 0.001) and significant directional pleiotropy (MR-Egger intercept (SE) = 0.02 (0.006), p < 0.001). Restricting the instrument to *cis*-variants completely attenuated the observed effect (OR (95% CI) 1.1 (0.52-2.34), p = 0.8) suggesting the association is driven by horizontal pleiotropy rather than a direct causal effect of CRP.

On reverse MR there was evidence for potentially causal effect of postoperative delirium on lower baseline CRP level, but the effect size was not consistent with clinically meaningful effects (OR (95% CI) 0.98 (0.97–0.99), p = 0.003). No significant evidence of reverse causation was identified for the other postoperative complication-inflammatory marker pairs assessed (Supplementary Table 3).

### Inflammation and all-cause outcomes: results from secondary analyses

The protective effect of genetically proxied IL-6 receptor blockade against stroke and MI seen in the primary postoperative analysis was substantially attenuated in the all-cause analysis (Supplementary Table 4). The Swerdlow et al. instrument no longer demonstrated an effect on stroke or MI, though the single-SNP Sarwar et al instrument showed a weak, suggestive protective effect against all-cause MI (Wald ratio OR (95% CI) 0.92 (0.85-1.00), p = 0.049) (Figure 2).

**Figure 2.**
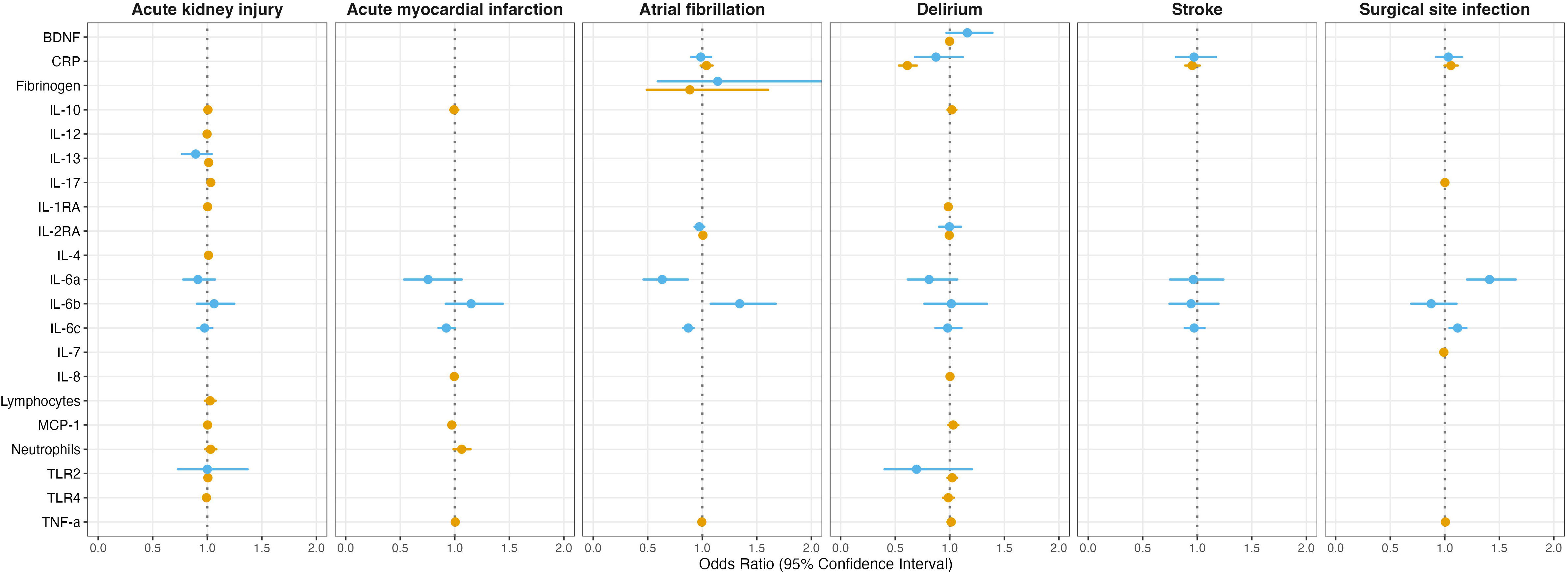
Odds ratios with 95% CIs from secondary Mendelian randomization analysis showing potential causal effects of genetically predicted levels of inflammatory markers on the risks of all-cause outcomes. Orange dots = trans variants, blue dots = cis variants. BDNF, brain-derived neurotrophic factor; CRP, C-reactive protein; IL, interleukin; IL6a, Swerdlow et al instrument; IL6b, Georgakis et al. instrument; IL-6c, Sarwar et al. instrument; MCP, monocyte chemoattractant protein; TLR, toll-like receptor; TNF, tumour necrosis factor. X-axes are capped at 2.0; where the upper 95% CI limit was beyond this point the line extends to the edge of the panel.

In contrast to the stroke and MI findings, strong evidence was found for causal effects of IL-6 signalling on all-cause AF across all three proxy instruments (Figure 2, Supplementary Table 4). The Sarwar et al. and Swerdlow et al. instruments were both associated with reduced risk of all-cause AF (OR (95% CI) 0.87 (0.82-0.92), p < 0.001 and 0.63 (0.46-0.87), p = 0.005 respectively). The Georgakis et al. instrument, which acts on IL-6 pathway activity in the opposite direction, was associated with increased risk of all-cause AF (OR 95% CI 1.34 (1.08-1.67), p = 0.009). Both multi-SNP instruments demonstrated significant heterogeneity on sensitivity analyses (Cochran’s Q = 12.6, p = 0.002 for Swerdlow et al. and 17.8, p = 0.007 for Georgakis et al.) though neither showed evidence of horizontal pleiotropy (MR-Egger intercept (SE) −0.07 (0.11), p = 0.63, and −0.02 (0.02), p = 0.4 respectively).

An additional association was identified between genetically proxied IL-6 and an increased risk of all-cause skin and subcutaneous tissue infection (the matched phenotype to SSI). This was present for the Swerdlow et al. and Sarwar et al. instruments (OR (95% CI) 1.41 (1.2-1.65), p < 0.001 and 1.12 (1.04-1.2), p = 0.002 respectively; Figure 2) with no evidence of instrument heterogeneity (Cochran’s Q = 0.12, p = 0.94) or horizontal pleiotropy (MR-Egger intercept (SE) 0.01 (0.05), p = 0.84) for the multi-SNP Swerdlow et al. instrument.

Results for the analysis of CRP on all-cause delirium mirrored those seen for postoperative delirium: the genome-wide analysis suggested a protective association (OR (95% CI) 0.61 (0.53-0.7), p < 0.001) but this was accompanied by severe heterogeneity (Cochran’s Q = 720, p < 0.001) and significant horizontal pleiotropy (MR-Egger intercept (SE) = 0.02 (0.003), p < 0.001). The effect was completely attenuated on *cis*-restricted analysis (OR (95% CI) 0.87 (0.68-1.12), p = 0.29; Figure 2) suggesting this was not a direct causal effect.

No significant evidence of reverse causation was seen on reverse MR for any of the exposure-outcome pairs analysed (Supplementary Table 5).

## Discussion

In this two-sample bi-directional Mendelian randomization study we investigated the potential causal relationship between a broad range of inflammatory markers and six common postoperative complications, as well as their all-cause equivalents. Our primary analysis of postoperative complications returned largely null results, except for suggestive protective effects of IL-6 signalling on cardiovascular outcomes. These results differed from those seen in the all-cause secondary analysis, suggesting that the role of inflammatory pathways in the acute postoperative period may be distinct from the effects seen in chronic disease pathology.

In the primary analysis, the Swerdlow et al. IL-6 instrument showed evidence of protective effects against postoperative stroke, with suggestive evidence of reduced risk of other cardiovascular outcomes (MI and AF) that was mirrored by the Sarwar et al. instrument. Whilst the variants in these instruments are associated with higher circulating levels of IL-6 and soluble IL-6R, they actually impair downstream inflammatory signalling, as evidenced by lower levels of C-reactive protein (CRP)^42,43^. This interpretation is further supported by our results using the Georgakis et al instrument, where variants associated with increased CRP (reflecting increased IL-6 signalling) showed point estimates of increased risk of stroke, AF and MI. These findings are consistent with the known role of IL-6 as a potent driver of endothelial activation and a pro-thrombotic state ^44^. In the acute perioperative period, individuals with genetically-proxied impairment of this pathway may therefore be relatively protected from the acute endothelial dysfunction and inflammatory cascade that precipitates perioperative stroke ^12^.

The divergence seen in the all-cause analysis underscores the complexity of this pathway in the perioperative setting. The protective effects on postoperative stroke and MI were largely absent in the all-cause analysis, whereas for AF we found consistent and robust protective effects across all three instruments studied, aligning with previous studies^15^. These results point to a role for IL-6 signalling in the pathogenesis of cardio- and cerebrovascular disease, though the contrast between postoperative and all-cause outcomes may suggest that the acute inflammatory insult of surgery is related to IL-6 mediated vascular events in a way that is distinct from chronic, low-grade inflammation over the life course. Furthermore, whilst our MR results for delirium were null, IL-6-mediated endothelial disruption remains a theoretical link between systemic inflammation and central complications. Current neuroinflammatory models of delirium implicate IL-6 in the disruption of the blood-brain barrier (BBB), which allows for the infiltration of peripheral inflammatory mediators into the central nervous system ^10^. The lack of a robust causal signal may indicate that the transient, acute-phase disruption of the BBB in the perioperative period is not captured by lifelong genetic proxies of baseline IL-6 signalling.

Our results estimating the effect of CRP on delirium were consistent across the postoperative and all-cause analyses, with genome-wide protective effects that were entirely abolished when restricted to *cis*-acting variants. This suggests that previous observational associations between CRP and delirium are likely driven by horizontal pleiotropy, where shared upstream pathways influence both the exposure and the outcome, rather than a direct causal effect of the circulating protein. This supports the concept of CRP as a biomarker rather than a causative factor, as has been reported with other outcome phenotypes ^19^, and reinforces the importance of *cis*-MR in inflammation analyses ^45^.

The lack of robust causal signals in the primary postoperative analysis, particularly for effects clearly observed in the all-cause analysis (e.g. IL-6 in AF), warrants further consideration. The genetic instruments used act as proxies for lifelong genetically-predicted baseline levels and do not necessarily reflect the inflammatory reactivity of an individual in the acute perioperative period. This temporal disconnect raises important questions regarding the biological interpretation of MR in this context: are we capturing an individual’s inherent baseline vulnerability to inflammation, the dynamic nature of their immune response to the stimulus of major surgery, or a combination of the two. Our null findings may suggest that for acute complications, the immediate perioperative reactivity of the system is more clinically relevant than lifelong genetic predisposition. This approach may therefore be less effective at modelling short-term outcomes in response to external stimuli compared with chronic disease onset resulting from lifelong exposure.

Furthermore, these null findings may arise from misspecification at the level of the mediator, pathway or biological scale. For markers such as CRP, the lack of a causal signal supports the interpretation that it functions primarily as an acute-phase reactant, reflecting the severity of physiological stress or disease activity, rather than acting as a direct causal driver of the complication itself. This is consistent with the neuroinflammatory model of delirium ^10^, in which central inflammatory processes may not be fully captured by systemic markers in the circulation. Circulating biomarkers may also fail to index local tissue inflammatory responses or the endorgan condition. The relative pathogenicity of a circulating cytokine may be contextdependent: factors such as advanced age or chronic comorbidities might lower the threshold for complications, meaning the genetically-predicted level of inflammation per se is not the only driver of adverse events in a vulnerable substrate. More broadly, this raises the possibility that the causal signals for these complications are not captured in their entirety by the specific mediators studied, or they operate at a biological scale or timepoint not captured by baseline protein levels.

The strengths of this study include the use of a bidirectional, two-sample MR design, which provides a robust framework for causal inference. Whilst there is a wealth of observational literature reporting associations between inflammatory markers and postoperative complications ^9, 46–49^, these associations are susceptible to confounding and reverse causation. Our study minimises these potential sources of bias by using genetic proxies and suggests that the inflammatory hypothesis of these complications is not simply a result of elevated levels of circulating cytokines.

However, there are limitations. The outcome GWAS in UK Biobank had a limited sample size for specific postoperative complications (for example delirium, approximately 1,000 cases), so the study was likely underpowered to detect small or
modest effects in the acute setting. This is compounded by the fact that the diagnoses relied upon ICD-10 clinical coding rather than prospective screening, a particular issue for delirium which may have a subclinical presentation. Hospital
coding for delirium is consistently found to have low sensitivity but excellent specificity ^50^ so whilst the source GWAS may have missed some cases any resulting bias would have reduced the observed effect. It is also possible, due to the nature of Hospital Episode Statistics data, that diagnoses may have pre-dated surgery.

However, the inclusion/exclusion criteria of the source GWAS took multiple steps to mitigate against this risk (Supplementary Methods 1). Similarly, the source GWAS for several exposures was Ahola-Olli et al. ^51^, which has a relatively small sample size and may have resulted in weak instrument bias (median F-statistics 16.35–17.29; minimum 4.36). Within MR it is possible for genetic variants to affect outcomes via alternative pathways (horizontal pleiotropy). To minimise this potential bias we prioritised *cis*-acting variants where possible and performed sensitivity analyses including Egger intercept. Our study is also potentially limited by the temporal disconnect between lifelong genetic proxies and acute surgical stimuli, as well as the potential for systemic markers to under-represent local tissue response and the role of vulnerable end-organ substrate, as described above. Our results therefore do not support the broad application of systemic immunomodulatory therapies in the prevention or treatment of the postoperative complications studied.

In summary, this study does not support a causal role for circulating inflammatory markers in the development of major postoperative complications. The observed associations in previous observational studies may therefore be the result of residual confounding or reflect the role of inflammatory markers as proxies for broader physiological stress. Further work should focus on the inflammatory response in the immediate perioperative context or the local interaction between inflammatory mediators and vulnerable end-organ substrate.

## Authors’ contributions

RAA: study design, data analysis, preparing first and subsequent drafts of the paper; PY: study design, drafting paper; BG: study design, drafting paper; GK: study design, drafting paper; TG: study design, drafting paper.

## Data availability statement

Summary statistics from the all-cause GWAS will be available from the University of Bristol data repository, data.bris, on publication.

Postoperative GWAS summary statistics are available from the University of Bristol data repository, data.bris, at https://doi.org/10.5523/bris.1m83zai2e26yq2lro3tixz9kqq (delirium) and https://doi.org/10.5523/bris.2qwk57ww5i25420f0sm64r7r3f (atrial fibrillation, acute kidney injury, myocardial infarction, stroke and surgical site infection).

## Acknowledgements

This research has been conducted using the UK Biobank Resource under Application Number 128619. This includes linked data provided by patients and collected by the NHS as part of their care and support and data assets made available by National Safe Haven as part of the Data and Connectivity National Core Study, led by Health Data Research UK in partnership with the Office for National Statistics and funded by UK Research and Innovation (grant ref MC_PC_20058).

Quality Control filtering of the UK Biobank data used the process described by R.Mitchell, G.Hemani, T.Dudding, L.Corbin, S.Harrison, L.Paternoster.

This work was carried out using the computational and data storage facilities of the Advanced Computing Research Centre, University of Bristol (http://www.bristol.ac.uk/acrc/).

