## Supplementary material for "Inflammation and postoperative complications after major surgery: a bi-directional two-sample Mendelian randomization study"

### STROBE-MR checklist of recommended items to address in reports of Mendelian randomization studies^1^ ^2^

| **Item No.** | **Section** | **Checklist item** | **Page No.** | **Relevant text from manuscript** |
| --- | --- | --- | --- | --- |
| 1 | **TITLE and ABSTRACT** | Indicate Mendelian randomization (MR) as the study’s design in the title and/or the abstract if that is a main purpose of the study | 1, 2 | Title: Inflammation and postoperative complications after major surgery: a bi-directional two-sample Mendelian randomization study  Abstract: We performed bi-directional two-sample Mendelian randomization (MR) to investigate… |
|  | **INTRODUCTION** |  |  |  |
| 2 | **Background** | Explain the scientific background and rationale for the reported study. What is the exposure? Is a potential causal relationship between exposure and outcome plausible? Justify why MR is a helpful method to address the study question | 4-5 | Throughout introduction |
| 3 | **Objectives** | State specific objectives clearly, including pre-specified causal hypotheses (if any). State that MR is a method that, under specific assumptions, intends to estimate causal effects | 4, 5 | Mendelian randomization (MR) is one approach to address the causal hypothesis by using genetic variants as proxies for the exposure of interest. Compared to other observational methods, MR is less susceptible to unmeasured confounding and reverse causation as genetic variants are randomly assigned and fixed at conception.  We undertook a two-sample MR study to systematically evaluate possible causal effects of genetically predicted circulating levels of inflammatory markers on the risk of six major postoperative complications: AF, AKI, AMI, delirium, stroke and SSI. |
|  | **METHODS** |  |  |  |
| 4 | **Study design and data sources** | Present key elements of the study design early in the article. Consider including a table listing sources of data for all phases of the study. For each data source contributing to the analysis, describe the following: |  |  |
|  | a) | Setting: Describe the study design and the underlying population, if possible. Describe the setting, locations, and relevant dates, including periods of recruitment, exposure, follow-up, and data collection, when available. | 6-7  Table 1 | Genetic variants were identified from summary statistics of large GWAS of European participants.  We used summary statistics from case-control GWAS of postoperative AF, AKI, AMI, delirium, SSI and stroke performed in UK Biobank [42,43]. |
|  | b) | Participants: Give the eligibility criteria, and the sources and methods of selection of participants. Report the sample size, and whether any power or sample size calculations were carried out prior to the main analysis | 6-7  Table 1, 2 | Participants were included if they underwent major surgery and had an ICD-10 coded complication of interest within 30 days of the procedure. Controls were those undergoing major surgery but not experiencing the complication.  To explore the relationship between postoperative complications and non-postoperative equivalent phenotypes (e.g. postoperative AF and chronic AF) we undertook a parallel analysis including all cases of each ICD-10 phenotype in the UK Biobank cohort (case-control numbers can be found in Supplementary Methods 2). For surgical site infection, an alternative diagnosis of ‘infections of the skin and subcutaneous tissue’ (ICD-10 codes L00-L08) was used. |
|  | c) | Describe measurement, quality control and selection of genetic variants | 6 | Genetic variants were identified from summary statistics of large GWAS of European participants. All variants associated with the outcome trait at a p value threshold of p < 10-5 and independent of each other with a clumping window of 10,000kb pairs and r2 threshold of < 0.001 were extracted as genome-wide/trans variants. Where present, cis variants were defined as those within a 1Mb window of the protein coding gene region based on Genome Reference Consortium Human Build 37. Cis-variants were considered the primary instruments for testing specific causal hypotheses for individual biomarkers. Trans-variants were included as supportive evidence to characterise the broader genetic architecture of the inflammatory response, acknowledging that these may act through indirect pathways (horizontal pleiotropy). Where a SNP was not present in the outcome dataset, proxy SNPs were identified using linkage disequilibrium (LD) tagging (r2 > 0.8) (LDlinkR [34]). |
|  | d) | For each exposure, outcome, and other relevant variables, describe methods of assessment and diagnostic criteria for diseases | 6-7 | Participants were included if they underwent major surgery and had an ICD-10 coded complication of interest within 30 days of the procedure. Controls were those undergoing major surgery but not experiencing the complication.  To explore the relationship between postoperative complications and non-postoperative equivalent phenotypes (e.g. postoperative AF and chronic AF) we undertook a parallel analysis including all cases of each ICD-10 phenotype in the UK Biobank cohort (case-control numbers can be found in Supplementary Methods 2). For surgical site infection, an alternative diagnosis of ‘infections of the skin and subcutaneous tissue’ (ICD-10 codes L00-L08) was used. Case-control GWAS adjusted for age, sex, genetic chip and the first ten ancestry principal components was performed for each outcome using regenie (v4.1 [44]). |
|  | e) | Provide details of ethics committee approval and participant informed consent, if relevant | 7 | The UK Biobank study was approved by the North-West Multi-centre Research Ethics Committee and all participants provided written informed consent. This research has been conducted under UK Biobank project number 128619. |
| 5 | **Assumptions** | Explicitly state the three core IV assumptions for the main analysis (relevance, independence and exclusion restriction) as well assumptions for any additional or sensitivity analysis | 6 | Mendelian randomization (MR) uses genetic variants associated with exposures and outcomes as proxies. For an instrument to be valid it must be: 1) reliably associated with the exposure (relevance assumption); 2) associated with the outcome only through the exposure (independence assumption); 3) independent of unobserved confounders that influence both exposure and outcome (exclusion-restriction) [34]. |
| 6 | **Statistical methods: main analysis** | Describe statistical methods and statistics used | 7-8 | Statistical analysis |
|  | a) | Describe how quantitative variables were handled in the analyses (i.e., scale, units, model) | N/A |  |
|  | b) | Describe how genetic variants were handled in the analyses and, if applicable, how their weights were selected | 6 | All variants associated with the outcome trait at a p value threshold of p < 10-5 and independent of each other with a clumping window of 10,000kb pairs and r2 threshold of < 0.001 were extracted as genome-wide/trans variants. Where present, cis variants were defined as those within a 1Mb window of the protein coding gene region based on Genome Reference Consortium Human Build 37. |
|  | c) | Describe the MR estimator (e.g. two-stage least squares, Wald ratio) and related statistics. Detail the included covariates and, in case of two-sample MR, whether the same covariate set was used for adjustment in the two samples | 7 | Our primary analysis was inverse-variance weighted (IVW) regression if >1 SNP was available or Wald ratio if the instrument comprised only 1 SNP. |
|  | d) | Explain how missing data were addressed | N/A |  |
|  | e) | If applicable, indicate how multiple testing was addressed | 7-8 | Statistical significance thresholds were adjusted for multiple testing using the Bonferroni method according to the number of exposures for each outcome (only those inflammatory markers reported to be associated with each outcome were analysed, Supplementary Table 1). |
| 7 | **Assessment of assumptions** | Describe any methods or prior knowledge used to assess the assumptions or justify their validity | 6 | The inflammatory markers included in the study were selected due to reported association with at least one of the postoperative complications of interest (Table 1, Supplementary Table 1). |
| 8 | **Sensitivity analyses and additional analyses** | Describe any sensitivity analyses or additional analyses performed (e.g. comparison of effect estimates from different approaches, independent replication, bias analytic techniques, validation of instruments, simulations) | 7 | Sensitivity analyses included MR-Egger, weighted median, weighted mode and MR Pleiotropy RESidual Sum and Outlier (MR-PRESSO, where outliers were identified). Where these methods required a minimum number of SNPs, they are reported when sufficient SNPs were available. Additionally, Steiger filtering, heterogeneity (Cochran’s Q-statistic) and pleiotropy (Egger intercept) were assessed. |
| 9 | **Software and pre-registration** |  |  |  |
|  | a) | Name statistical software and package(s), including version and settings used | 7 | Bidirectional two-sample MR was performed using the TwoSampleMR package (v. 0.6.25 [45,46]) in R (4.5.0). |
|  | b) | State whether the study protocol and details were pre-registered (as well as when and where) | N/A |  |
|  | **RESULTS** |  |  |  |
| 10 | **Descriptive data** |  |  |  |
|  | a) | Report the numbers of individuals at each stage of included studies and reasons for exclusion. Consider use of a flow diagram |  | Tables 1 and 2 |
|  | b) | Report summary statistics for phenotypic exposure(s), outcome(s), and other relevant variables (e.g. means, SDs, proportions) | N/A |  |
|  | c) | If the data sources include meta-analyses of previous studies, provide the assessments of heterogeneity across these studies | N/A |  |
|  | d) | For two-sample MR:  i.  Provide justification of the similarity of the genetic variant-exposure associations between the exposure and outcome samples  ii.  Provide information on the number of individuals who overlap between the exposure and outcome studies | 7 | Exposure and outcome GWAS were from different consortia, minimising the risk of bias from sample overlap. |
| 11 | **Main results** |  |  |  |
|  | a) | Report the associations between genetic variant and exposure, and between genetic variant and outcome, preferably on an interpretable scale | 9-11 | Throughout Results |
|  | b) | Report MR estimates of the relationship between exposure and outcome, and the measures of uncertainty from the MR analysis, on an interpretable scale, such as odds ratio or relative risk per SD difference | 9-11 | Throughout Results |
|  | c) | If relevant, consider translating estimates of relative risk into absolute risk for a meaningful time period | N/A |  |
|  | d) | Consider plots to visualize results (e.g. forest plot, scatterplot of associations between genetic variants and outcome versus between genetic variants and exposure) |  | Figure 1, Figure 2 |
| 12 | **Assessment of assumptions** |  |  |  |
|  | a) | Report the assessment of the validity of the assumptions | 9-11 | Throughout Results |
|  | b) | Report any additional statistics (e.g., assessments of heterogeneity across genetic variants, such as *I^2^*, Q statistic or E-value) | 9-11 | Throughout Results |
| 13 | **Sensitivity analyses and additional analyses** |  |  |  |
|  | a) | Report any sensitivity analyses to assess the robustness of the main results to violations of the assumptions | 9-11 | Throughout Results |
|  | b) | Report results from other sensitivity analyses or additional analyses | 9-11 | Throughout Results |
|  | c) | Report any assessment of direction of causal relationship (e.g., bidirectional MR) | 9-11 | Throughout Results |
|  | d) | When relevant, report and compare with estimates from non-MR analyses | N/A |  |
|  | e) | Consider additional plots to visualize results (e.g., leave-one-out analyses) | N/A |  |
|  | **DISCUSSION** |  |  |  |
| 14 | **Key results** | Summarize key results with reference to study objectives | 12 | Our primary analysis of postoperative complications returned largely null results, except for suggestive protective effects of IL-6 signalling on cardiovascular outcomes. These results differed from those seen in the all-cause secondary analysis, suggesting that the role of inflammatory pathways in the acute postoperative period may be distinct from the effects seen in chronic disease pathology. |
| 15 | **Limitations** | Discuss limitations of the study, taking into account the validity of the IV assumptions, other sources of potential bias, and imprecision. Discuss both direction and magnitude of any potential bias and any efforts to address them | 13-14 | Limitations paragraph |
| 16 | **Interpretation** |  |  |  |
|  | a) | Meaning: Give a cautious overall interpretation of results in the context of their limitations and in comparison with other studies | 12, 14 | … suggesting that the role of inflammatory pathways in the acute postoperative period may be distinct from the effects seen in chronic disease pathology.  In summary, this study does not support a causal role for circulating inflammatory markers in the development of major postoperative complications. The observed associations in previous observational studies may therefore be the result of residual confounding or reflect the role of inflammatory markers as proxies for broader physiological stress. Further work should focus on the inflammatory response in the immediate perioperative context or the local interaction between inflammatory mediators and vulnerable end-organ substrate. |
|  | b) | Mechanism: Discuss underlying biological mechanisms that could drive a potential causal relationship between the investigated exposure and the outcome, and whether the gene-environment equivalence assumption is reasonable. Use causal language carefully, clarifying that IV estimates may provide causal effects only under certain assumptions | 12, 13 | These results point to a role for IL-6 in the pathogenesis of cardio- and cerebrovascular disease, though the divergence between postoperative and all-cause outcomes may suggest that the acute inflammatory insult of surgery is related to IL-6 mediated vascular events in a way that is distinct from chronic, low-grade inflammation over the life course.  The genetic instruments used act as proxies for lifelong genetically-predicted baseline levels and do not necessarily reflect the inflammatory reactivity of an individual in the acute perioperative period. They may therefore be less effective at modelling short-term outcomes in response to the external stimulus of major surgery compared with chronic disease onset resulting from lifelong exposure.  The relative pathogenicity of a circulating cytokine may also depend upon the condition of the end-organ tissue: factors such as advanced age or chronic comorbidities might lower the threshold for complications, meaning the genetically-predicted level of inflammation per se is not the only driver of adverse events in vulnerable substrate. |
|  | c) | Clinical relevance: Discuss whether the results have clinical or public policy relevance, and to what extent they inform effect sizes of possible interventions | 12, 13-14 | These results point to a role for IL-6 in the pathogenesis of cardio- and cerebrovascular disease, though the divergence between postoperative and all-cause outcomes may suggest that the acute inflammatory insult of surgery is related to IL-6 mediated vascular events in a way that is distinct from chronic, low-grade inflammation over the life course.  The genetic instruments used act as proxies for lifelong genetically-predicted baseline levels and do not necessarily reflect the inflammatory reactivity of an individual in the acute perioperative period. They may therefore be less effective at modelling short-term outcomes in response to the external stimulus of major surgery compared with chronic disease onset resulting from lifelong exposure.  Our results therefore do not support the broad application of systemic immunomodulatory therapies in the prevention or treatment of the postoperative complications studied. |
| 17 | **Generalizability** | Discuss the generalizability of the study results (a) to other populations, (b) across other exposure periods/timings, and (c) across other levels of exposure | 13 | In addition to the temporal disconnect between acute postoperative events and the MR approach, as described above, systemic levels of inflammatory biomarkers may not capture local tissue inflammatory responses. The relative pathogenicity of a circulating cytokine may also depend upon the condition of the end-organ tissue: factors such as advanced age or chronic comorbidities might lower the threshold for complications, meaning the genetically-predicted level of inflammation per se is not the only driver of adverse events in vulnerable substrate |
|  | **OTHER INFORMATION** |  |  |  |
| 18 | **Funding** | Describe sources of funding and the role of funders in the present study and, if applicable, sources of funding for the databases and original study or studies on which the present study is based | 19 | Funding statement |
| 19 | **Data and data sharing** | Provide the data used to perform all analyses or report where and how the data can be accessed, and reference these sources in the article. Provide the statistical code needed to reproduce the results in the article, or report whether the code is publicly accessible and if so, where | Table 1  8, 16 | Citations given for all data in Table 1  Code to support the analyses is available on GitHub [48].  Data availability statement |
| 20 | **Conflicts of Interest** | All authors should declare all potential conflicts of interest | 18 | Declaration of interests statement |

This checklist is copyrighted by the Equator Network under the Creative Commons Attribution 3.0 Unported (CC BY 3.0) license.

1.	Skrivankova VW, Richmond RC, Woolf BAR, Yarmolinsky J, Davies NM, Swanson SA, et al. Strengthening the Reporting of Observational Studies in Epidemiology using Mendelian Randomization (STROBE-MR) Statement. JAMA. 2021;under review.

2.	Skrivankova VW, Richmond RC, Woolf BAR, Davies NM, Swanson SA, VanderWeele TJ, et al. Strengthening the Reporting of Observational Studies in Epidemiology using Mendelian Randomisation (STROBE-MR): Explanation and Elaboration. BMJ. 2021;375:n2233.

### Supplementary methods 1: Mendelian Randomization framework and assumptions

The following Directed acyclic graph (DAG) illustrates the assumptions of the Mendelian Randomization framework:

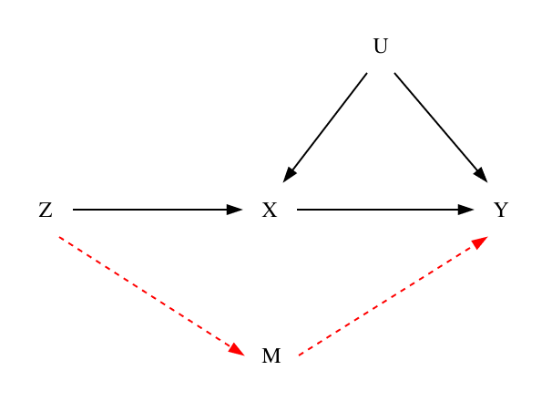

*Z*: genetic instrumental variables (inflammatory polymorphisms); *X*: exposure of interest (circulating inflammatory marker levels); *Y*: outcome (postoperative complications); *U*: unobserved postnatal confounders (e.g. lifestyle factors, comorbidities); *M*: pleiotropic pathway (representing ‘multiple diseases’ identified by the reviewer).

For an instrument to be valid it must be:

1. reliably associated with the exposure (relevance assumption, path Z -> X);
2. associated with the outcome only through the exposure (independence assumption; no path via M);
3. independent of unobserved confounders that influence both exposure and outcome (exclusion-restriction; no path from U to Z)

The potential causal path flows from genetic variants (Z), acting as instrumental variables for circulating inflammatory markers (X), to the postoperative outcome (Y). As shown in the DAG, genetic variants are inherently independent of postnatal unobserved confounders (U) as they are randomly assigned and fixed at conception. This ‘independence assumption’ is represented by the absence of a path from U to Z.

The path Z -> M -> Y represents potential horizontal pleiotropy, where the genetic variant influences the outcome through pathways other than the exposure of interest (e.g. comorbidities). We addressed the risk of horizontal pleiotropy by:

- prioritising cis-restricted analyses: variants biologically anchored to the specific protein coding region are less likely to have pleiotropic effects via other pathways (M)
- sensitivity analyses: we used MR-Egger regression to detect and quantify directional pleiotropy..

Finally, adjusting for comorbidities (M) in an MR framework risks introducing collider bias. If the comorbidity is influenced by both the genetic instrument (Z) and unmeasured confounders (U), conditioning on M can create a spurious association between Z and U, thereby invalidating the causal estimate.

### Supplementary methods 2: Case definition, inclusion and exclusion criteria for postoperative outcome GWAS

The following provides additional detail on the methods of the source GWAS used for outcome data.

UK Biobank (UKB) contains linked Hospital Episode Statistics (HES) inpatient data for participants. Eligible participants were those undergoing major, inpatient surgery (as defined below) after their date of enrolment in UK Biobank. Exclusion criteria for all phenotypes included a previous diagnosis of the outcome of interest and planned day case surgery. Individuals with a previous diagnosis of the outcome phenotype of interest were excluded to reduce potential bias through confounding and reverse causation. It also ensured all events occurred after baseline variables were measured at UKB enrolment and contributed to a more homogenous case-control cohort. For acute MI, cardiothoracic and cardiology procedures were excluded to reduce the risk of the diagnosis preceding the procedure. Cases were identified according to ICD-10 diagnosis code(s), using specific codes for acute diagnoses where possible:

| **Code** | **Diagnosis** | **Notes** |
| --- | --- | --- |
| F05 | Delirium | Delirium is an acute event by definition |
| I48.9 | Atrial fibrillation and flutter, unspecified | This code was chosen as it is distinct from chronic diagnosis codes, e.g. I48.2 Chronic atrial fibrillation |
| N17 | Acute renal failure | Acute by definition |
| I61/63/64 | Intracerebral haemorrhage / Cerebral infarction / Stroke, not specified as haemorrhage or infarction |  |
| T81.4 | Infection following a procedure, not elsewhere classified | A specific postprocedural code |

The operation table contains details of all OPCS4 procedure codes for a patient during a given admission. Inpatient surgery was defined according to patient class (excluding day case). Major surgery was defined by using a combination of two grading systems from the literature (Bupa schedule of procedures(1) and Abbott(2)). Eligible procedures were either Bupa ‘major’ or ‘complex’ category or, if not classified in Bupa, in the ‘restrictive’ category as defined by Abbott. Only procedures performed after enrolment in UKB were eligible. All surgical specialties were included except for the acute myocardial infarction cohort, where cardiothoracic and cardiology procedures were excluded.

In cases of multiple procedures on the same date, the procedure coded as level 1 was treated as the index procedure. If there were multiple or no level 1 procedures, the procedure with the highest grade was used (complex > major > restrictive). If there were multiple procedures with the highest grade, minimum array index was used as a tiebreak. Count variables were added to account for a) multiple surgical procedures within the same hospital admission and b) cumulative procedures since UKB enrolment.

A limitation of HES coding is that procedures have a specific date of operation, whereas diagnoses are linked to an entire inpatient episode (without a specific date of diagnosis). To limit the risk of a diagnosis preceding an operative procedure where both occurred in the same inpatient episode, the following decision tree was used:

Complication diagnosis in later episode than operation date?

Yes: include

No

Operation and diagnosis in same episode. Time from episode start to operation <14 days?

Yes

Operation date in first half of episode?

Yes: include

No: exclude

No: exclude

### Supplementary methods 3: Case-control numbers for all-cause GWAS in UK Biobank

| **Complication** | **ICD-10 diagnosis code** | **Cases** | **Controls** |
| --- | --- | --- | --- |
| Atrial fibrillation | I48.9 | 35,103 | 451,256 |
| Acute kidney injury | N17 | 21,175 | 465,184 |
| Acute myocardial infarction | I21 | 15,719 | 470,640 |
| Delirium | F05 | 6,556 | 479,803 |
| Stroke | I61, I63, I64 | 11,410 | 474,949 |
| infections of the skin and subcutaneous tissue | L00-L08 | 20,697 | 465,662 |

### Supplementary table 1: Exposures analysed for each outcome, citations for observational associations and Bonferroni-adjusted p-value thresholds

|  | **Postoperative complication (outcome)** | | | | | |
| --- | --- | --- | --- | --- | --- | --- |
| **Inflammatory marker (exposure)** | **AF** | **AKI** | **Delirium** | **AMI** | **SSI** | **Stroke** |
| BDNF |  |  | X [1,2] |  |  |  |
| CRP | X [3] |  | X [2,4,5] |  | X [6] | X [7] |
| Fibrinogen | X [3,8,9] |  |  |  |  |  |
| IL-1 |  | X [10] | X [1,2,11] | X [12] |  |  |
| IL-2 | X [3] |  | X [13] |  |  |  |
| IL-4 |  | X [10] |  |  |  |  |
| IL-6 | X [3,8] | X [10] | X [1,2,4,13–15] | X [12] | X [16] | X [7] |
| IL-7 |  |  |  |  | X [17] |  |
| IL-8 |  |  | X [1,2,4,14,15] | X [12] |  |  |
| IL-10 |  | X [18] | X [1,2,4,14] | X [12] |  |  |
| IL-12 |  | X [10] |  |  |  |  |
| IL-13 |  | X [10] |  |  |  |  |
| IL-17 |  | X [10] |  |  | X [17] |  |
| Lymphocytes |  | X [10] |  |  |  |  |
| MCP1 |  | X [10] | X [1] | X [12] |  |  |
| Neutrophils |  | X [10,18] |  | X [12] |  |  |
| TLR2 |  | X [10,18] | X [2] |  |  |  |
| TLR4 |  | X [10] | X [1,2,14] |  |  |  |
| TNF-a | X [3,9] | X [10] | X [1,2,4,11,14,15,19] | X [12] | X [16] |  |
| **Total** | 5 | 13 | 11 | 7 | 5 | 2 |
| **Bonferroni-adjusted p value** | **0.01** | **0.0038** | **0.0045** | **0.0071** | **0.01** | **0.025** |

### Supplementary Table 2: Results of primary analysis. Odds ratios with 95% CIs from Mendelian randomization analysis showing associations between genetically predicted levels of inflammatory markers and risks for postoperative complications. IL-6a: Swerdlow et al; IL-6b: Georgakis et al; IL-6c: Sarwar et al.

| Outcome | Exposure | Analysis | n SNPs | Odds ratio (95% CI) | P-value |
| --- | --- | --- | --- | --- | --- |
| Acute kidney injury | IL-10 | Trans | 113 | 0.97 (0.91–1.03) | 0.38 |
|  | IL-12 | Trans | 118 | 0.98 (0.92–1.04) | 0.56 |
|  | IL-13 | Trans | 110 | 1.03 (0.99–1.08) | 0.22 |
|  | IL-13 | Cis | 1 | 1.27 (0.86–1.89) | 0.23 |
|  | IL-17 | Trans | 94 | 1.03 (0.96–1.10) | 0.32 |
|  | IL-1RA | Trans | 106 | 1.01 (0.96–1.06) | 0.72 |
|  | IL-4 | Trans | 100 | 1.01 (0.94–1.08) | 0.70 |
|  | IL-6a | Cis | 3 | 1.15 (0.76–1.74) | 0.51 |
|  | IL-6b | Cis | 7 | 1.03 (0.68–1.56) | 0.87 |
|  | IL-6c | Cis | 1 | 1.06 (0.88–1.27) | 0.50 |
|  | Lymphocytes | Trans | 421 | 1.09 (0.98–1.23) | 0.14 |
|  | MCP-1 | Trans | 94 | 1.04 (0.97–1.11) | 0.25 |
|  | Neutrophils | Trans | 402 | 1.03 (0.91–1.16) | 0.58 |
|  | TLR2 | Trans | 159 | 0.95 (0.88–1.03) | 0.23 |
|  | TLR2 | Cis | 1 | 1.15 (0.51–2.61) | 0.74 |
|  | TLR4 | Trans | 151 | 0.99 (0.92–1.07) | 0.84 |
| Acute myocardial infarction | IL-10 | Trans | 113 | 0.87 (0.71–1.07) | 0.18 |
|  | IL-6a | Cis | 3 | 0.24 (0.06–0.90) | 0.04 |
|  | IL-6b | Cis | 7 | 2.23 (0.59–8.43) | 0.24 |
|  | IL-6c | Cis | 1 | 0.75 (0.42–1.34) | 0.32 |
|  | IL-8 | Trans | 99 | 0.94 (0.82–1.09) | 0.39 |
|  | MCP-1 | Trans | 94 | 0.85 (0.69–1.05) | 0.14 |
|  | Neutrophils | Trans | 402 | 1.23 (0.84–1.81) | 0.28 |
|  | TNF-a | Trans | 96 | 0.97 (0.84–1.13) | 0.70 |
| Atrial fibrillation | CRP | Trans | 249 | 0.97 (0.85–1.11) | 0.67 |
|  | CRP | Cis | 3 | 0.89 (0.65–1.21) | 0.43 |
|  | Fibrinogen | Trans | 29 | 1.46 (0.40–5.30) | 0.56 |
|  | Fibrinogen | Cis | 1 | 0.86 (0.09–8.50) | 0.89 |
|  | IL-2RA | Trans | 212 | 0.99 (0.94–1.04) | 0.69 |
|  | IL-2RA | Cis | 1 | 0.97 (0.83–1.13) | 0.68 |
|  | IL-6a | Cis | 3 | 0.59 (0.39–0.91) | 0.02 |
|  | IL-6b | Cis | 7 | 1.14 (0.73–1.78) | 0.58 |
|  | IL-6c | Cis | 1 | 0.87 (0.72–1.05) | 0.13 |
|  | TNF-a | Trans | 96 | 0.99 (0.94–1.04) | 0.70 |
| Delirium | BDNF | Trans | 179 | 0.96 (0.89–1.04) | 0.30 |
|  | BDNF | Cis | 4 | 0.9 (0.58–1.42) | 0.80 |
|  | CRP | Trans | 249 | 0.62 (0.49–0.78) | 5.1 x10^-5^ |
|  | CRP | Cis | 3 | 1.11 (0.52–2.34) | 0.8 |
|  | IL-10 | Trans | 113 | 1.04 (0.93–1.17) | 0.47 |
|  | IL-1RA | Trans | 106 | 0.99 (0.91–1.08) | 0.79 |
|  | IL-2RA | Trans | 212 | 1.02 (0.92–1.13) | 0.65 |
|  | IL-2RA | Cis | 1 | 1.22 (0.95–1.57) | 0.12 |
|  | IL-6a | Cis | 3 | 0.78 (0.20–3.08) | 0.73 |
|  | IL-6b | Cis | 7 | 0.82 (0.40–1.66) | 0.58 |
|  | IL-6c | Cis | 1 | 1.05 (0.77–1.43) | 0.77 |
|  | IL-8 | Trans | 99 | 1.08 (1.00–1.17) | 0.04 |
|  | MCP-1 | Trans | 94 | 1.08 (0.97–1.21) | 0.17 |
|  | TLR2 | Trans | 159 | 1.05 (0.93–1.19) | 0.42 |
|  | TLR2 | Cis | 1 | 0.73 (0.18–2.89) | 0.65 |
|  | TLR4 | Trans | 151 | 0.90 (0.80–1.03) | 0.12 |
|  | TNF-a | Trans | 96 | 1.02 (0.94–1.10) | 0.65 |
| Stroke | CRP | Trans | 249 | 0.84 (0.64–1.11) | 0.23 |
|  | CRP | Cis | 3 | 0.65 (0.33–1.29) | 0.21 |
|  | IL-6a | Cis | 3 | 0.27 (0.11–0.69) | 0.006 |
|  | IL-6b | Cis | 7 | 2.51 (0.98–6.45) | 0.06 |
|  | IL-6c | Cis | 1 | 0.68 (0.45–1.02) | 0.07 |
| Surgical site infection | CRP | Trans | 249 | 0.90 (0.78–1.02) | 0.09 |
|  | CRP | Cis | 3 | 0.7 (0.46–1.07) | 0.1 |
|  | IL-17 | Trans | 94 | 1.00 (0.93–1.08) | 0.95 |
|  | IL-6a | Cis | 3 | 1.12 (0.70–1.77) | 0.65 |
|  | IL-6b | Cis | 7 | 0.88 (0.57–1.36) | 0.56 |
|  | IL-6c | Cis | 1 | 1.02 (0.84–1.24) | 0.82 |
|  | IL-7 | Trans | 96 | 1.04 (0.99–1.10) | 0.11 |
|  | TNF-a | Trans | 96 | 1.01 (0.96–1.06) | 0.67 |

### Supplementary table 3: Results of reverse Mendelian randomisation analysis with inflammatory markers as outcome and postoperative complications as exposure

NB: For IL-6, Konieczny et al (<https://doi.org/10.1038/s42003-025-07453-w>) was used as the outcome GWAS as full summary statistics were not available for the IL-6 references used in the forward analysis. P-value threshold p<10^-4^ and clumped r^2^ < 0.001, 10000kB.

| **Outcome** | **Exposure** | **N SNPs** | **Odds ratio (95% CI)** | **P value** |
| --- | --- | --- | --- | --- |
| BDNF | Delirium | 2250 | 1.01 (1–1.02) | 0.06 |
| CRP | AF | 179 | 1 (0.99–1.01) | 0.12 |
|  | Delirium | 179 | 0.98 (0.97–0.99) | 0.003 |
|  | SSI | 179 | 0.99 (0.97–1.02) | 0.66 |
|  | Stroke | 179 | 0.99 (0.98–1) | 0.14 |
| IL-10 | AKI | 393 | 0.97 (0.92–1.03) | 0.33 |
|  | AMI | 393 | 1 (0.98–1.02) | 0.60 |
|  | Delirium | 393 | 1 (0.97–1.03) | 0.77 |
| IL-12(p70) | AKI | 389 | 0.96 (0.91–1.01) | 0.19 |
| IL-17 | AKI | 392 | 0.98 (0.93–1.04) | 0.52 |
|  | SSI | 392 | 1.01 (0.96–1.07) | 0.67 |
| IL-1RA | AKI | 381 | 0.98 (0.9–1.06) | 0.61 |
|  | Delirium | 381 | 0.97 (0.92–1.02) | 0.23 |
| IL-6 | AF | 2973 | 1 (1–1) | 0.64 |
|  | AMI | 2974 | 1 (1–1) | 0.80 |
|  | Delirium | 2974 | 1 (1–1) | 0.68 |
|  | SSI | 2974 | 1 (1–1) | 0.43 |
|  | Stroke | 2974 | 1 (1–1) | 0.20 |
| IL-7 | SSI | 379 | 0.98 (0.9–1.06) | 0.57 |
| IL-8 | AMI | 376 | 1 (0.97–1.03) | 0.86 |
|  | Delirium | 376 | 0.98 (0.93–1.03) | 0.54 |
| IL13 | AKI | 383 | 0.96 (0.89–1.04) | 0.38 |
| Lymphocytes | AKI | 566 | 1 (0.99–1.01) | 0.53 |
| MCP-1 | AKI | 389 | 0.97 (0.92–1.02) | 0.33 |
|  | AMI | 389 | 1 (0.98–1.02) | 0.65 |
|  | Delirium | 389 | 1 (0.97–1.03) | 0.88 |
| Neutrophils | AKI | 566 | 1 (0.99–1.01) | 0.51 |
|  | AMI | 566 | 1 (1–1) | 0.24 |
| sIL-2Ra | AF | 381 | 1 (0.98–1.02) | 0.78 |
|  | Delirium | 381 | 1 (0.95–1.05) | 0.89 |
| TLR2 | AKI | 2438 | 1 (0.99–1.01) | 0.53 |
|  | Delirium | 2438 | 1 (0.99–1.01) | 0.49 |
| TLR4 | AKI | 2438 | 1 (0.99–1.01) | 0.77 |
|  | Delirium | 2438 | 1 (0.99–1.01) | 0.22 |
| TNF-a | AF | 379 | 0.99 (0.97–1.01) | 0.57 |
|  | AMI | 379 | 1 (0.97–1.03) | 0.84 |
|  | Delirium | 379 | 0.98 (0.93–1.03) | 0.46 |
|  | SSI | 379 | 0.95 (0.88–1.03) | 0.23 |

### Supplementary Table 4: Results of secondary analysis. Odds ratios with 95% CIs from Mendelian randomization analysis showing associations between genetically predicted levels of inflammatory markers and risks for all-cause outcomes. IL-6a: Swerdlow et al; IL-6b: Georgakis et al; IL-6c: Sarwar et al.

| Outcome | Exposure | Analysis | n SNPs | Odds ratio (95% CI) | P-value |
| --- | --- | --- | --- | --- | --- |
| Acute kidney injury | IL-10 | Trans | 113 | 1.01 (0.98–1.03) | 0.63 |
|  | IL-12 | Trans | 118 | 1.01 (0.97–1.02) | 0.86 |
|  | IL-13 | Trans | 110 | 1.08 (0.77–1.04) | 0.15 |
|  | IL-13 | Cis | 1 | 1.01 (1.00–1.03) | 0.12 |
|  | IL-17 | Trans | 94 | 1.01 (1.01–1.06) | 0.02 |
|  | IL-1RA | Trans | 106 | 1.01 (0.98–1.02) | 0.73 |
|  | IL-4 | Trans | 100 | 1.01 (0.98–1.04) | 0.42 |
|  | IL-6a | Cis | 3 | 1.08 (0.78–1.07) | 0.27 |
|  | IL-6b | Cis | 7 | 1.09 (0.90–1.25) | 0.46 |
|  | IL-6c | Cis | 1 | 1.04 (0.91–1.05) | 0.48 |
|  | Lymphocytes | Trans | 421 | 1.02 (0.98–1.08) | 0.29 |
|  | MCP-1 | Trans | 94 | 1.02 (0.97–1.03) | 0.84 |
|  | Neutrophils | Trans | 402 | 1.03 (0.98–1.08) | 0.24 |
|  | TLR2 | Trans | 159 | 1.01 (0.98–1.03) | 0.71 |
|  | TLR2 | Cis | 1 | 1.17 (0.73–1.37) | 1.00 |
|  | TLR4 | Trans | 151 | 1.02 (0.96–1.02) | 0.61 |
| Acute myocardial infarction | IL-10 | Trans | 113 | 1.02 (0.96–1.03) | 0.75 |
|  | IL-6a | Cis | 3 | 1.19 (0.54–1.06) | 0.11 |
|  | IL-6b | Cis | 7 | 1.12 (0.92–1.44) | 0.23 |
|  | IL-6c | Cis | 1 | 1.04 (0.85–1.00) | 0.05 |
|  | IL-8 | Trans | 99 | 1.01 (0.98–1.02) | 0.62 |
|  | MCP-1 | Trans | 94 | 1.02 (0.94–1.01) | 0.12 |
|  | Neutrophils | Trans | 402 | 1.04 (0.99–1.15) | 0.11 |
|  | TNF-a | Trans | 96 | 1.01 (0.98–1.03) | 0.71 |
| Atrial fibrillation | CRP | Trans | 249 | 1.03 (0.98–1.10) | 0.16 |
|  | CRP | Cis | 3 | 1.05 (0.90–1.08) | 0.76 |
|  | Fibrinogen | Trans | 29 | 1.35 (0.49–1.60) | 0.69 |
|  | Fibrinogen | Cis | 1 | 1.40 (0.59–2.20) | 0.69 |
|  | IL-2RA | Trans | 212 | 1.01 (0.99–1.02) | 0.42 |
|  | IL-2RA | Cis | 1 | 1.02 (0.93–1.02) | 0.24 |
|  | IL-6a | Cis | 3 | 1.18 (0.46–0.87) | 0.005 |
|  | IL-6b | Cis | 7 | 1.12 (1.08–1.67) | 0.01 |
|  | IL-6c | Cis | 1 | 1.03 (0.82–0.92) | 1.4 x10 ^-6^ |
|  | TNF-a | Trans | 96 | 1.01 (0.98–1.01) | 0.56 |
| Delirium | BDNF | Trans | 179 | 1.02 (0.97–1.03) | 0.91 |
|  | BDNF | Cis | 4 | 1.10 (0.97–1.39) | 0.11 |
|  | CRP | Trans | 249 | 1.07 (0.53–0.70) | 6.7 x10^-13^ |
|  | CRP | Cis | 3 | 1.14 (0.68–1.12) | 0.29 |
|  | IL-10 | Trans | 113 | 1.02 (0.98–1.06) | 0.36 |
|  | IL-1RA | Trans | 106 | 1.02 (0.96–1.02) | 0.39 |
|  | IL-2RA | Trans | 212 | 1.02 (0.97–1.03) | 0.75 |
|  | IL-2RA | Cis | 1 | 1.05 (0.90–1.10) | 0.95 |
|  | IL-6a | Cis | 3 | 1.15 (0.61–1.07) | 0.14 |
|  | IL-6b | Cis | 7 | 1.15 (0.77–1.34) | 0.92 |
|  | IL-6c | Cis | 1 | 1.06 (0.87–1.11) | 0.76 |
|  | IL-8 | Trans | 99 | 1.02 (0.97–1.03) | 0.89 |
|  | MCP-1 | Trans | 94 | 1.02 (0.99–1.08) | 0.18 |
|  | TLR2 | Trans | 159 | 1.02 (0.98–1.07) | 0.32 |
|  | TLR2 | Cis | 1 | 1.32 (0.40–1.20) | 0.19 |
|  | TLR4 | Trans | 151 | 1.03 (0.94–1.04) | 0.60 |
|  | TNF-a | Trans | 96 | 1.02 (0.98–1.05) | 0.46 |
| Stroke | CRP | Trans | 249 | 1.04 (0.89–1.02) | 0.19 |
|  | CRP | Cis | 3 | 1.10 (0.80–1.17) | 0.75 |
|  | IL-6a | Cis | 3 | 1.14 (0.75–1.24) | 0.78 |
|  | IL-6b | Cis | 7 | 1.13 (0.74–1.20) | 0.63 |
|  | IL-6c | Cis | 1 | 1.05 (0.88–1.07) | 0.55 |
| Infections of the skin and subcutaneous tissue | CRP | Trans | 249 | 1.03 (1.00–1.12) | 0.06 |
|  | CRP | Cis | 3 | 1.06 (0.92–1.16) | 0.58 |
|  | IL-17 | Trans | 94 | 1.01 (0.97–1.03) | 0.92 |
|  | IL-6a | Cis | 3 | 1.08 (1.20–1.65) | 1.9 x10^-5^ |
|  | IL-6b | Cis | 7 | 1.13 (0.69–1.11) | 0.27 |
|  | IL-6c | Cis | 1 | 1.04 (1.04–1.20) | 0.002 |
|  | IL-7 | Trans | 96 | 1.01 (0.97–1.01) | 0.39 |
|  | TNF-a | Trans | 96 | 1.01 (0.99–1.02) | 0.52 |

### Supplementary table 5: Results of reverse Mendelian randomisation analysis with inflammatory markers as outcome and all-cause phenotypes as exposure

NB: For IL-6, Konieczny et al (<https://doi.org/10.1038/s42003-025-07453-w>) was used as the outcome GWAS as full summary statistics were not available for the IL-6 references used in the forward analysis. P-value threshold p<10^-4^ and clumped r^2^ < 0.001, 10000kB.

| **Outcome** | **Exposure** | **N SNPs** | **Odds ratio (95% CI)** | **P value** |
| --- | --- | --- | --- | --- |
| BDNF | Delirium | 2250 | 1.03 (0.99 –1.07) | 0.16 |
| CRP | AF | 179 | 1.01 (0.99 –1.03) | 0.58 |
|  | Delirium | 179 | 0.99 (0.96 –1.03) | 0.58 |
|  | SSI | 179 | 1.04 (0.98 –1.1) | 0.13 |
|  | Stroke | 179 | 1.02 (0.98 –1.06) | 0.30 |
| IL-10 | AKI | 393 | 0.96 (0.84 –1.1) | 0.54 |
|  | AMI | 393 | 1.02 (0.9 –1.15) | 0.80 |
|  | Delirium | 393 | 0.97 (0.89 –1.06) | 0.48 |
| IL-12(p70) | AKI | 389 | 1.01 (0.89 –1.15) | 0.90 |
| IL-17 | AKI | 392 | 0.99 (0.87 –1.13) | 0.86 |
|  | SSI | 392 | 1.05 (0.92 –1.2) | 0.48 |
| IL-1RA | AKI | 381 | 1.01 (0.83 –1.23) | 0.94 |
|  | Delirium | 381 | 0.95 (0.83 –1.08) | 0.46 |
| IL-6 | AF | 2973 | 1.01 (0.99 –1.03) | 0.51 |
|  | AMI | 2974 | 1.02 (1 –1.04) | 0.07 |
|  | Delirium | 2974 | 1 (0.99 –1.01) | 0.68 |
|  | SSI | 2974 | 1 (0.98 –1.02) | 0.80 |
|  | Stroke | 2974 | 1 (0.99 –1.01) | 0.52 |
| IL-7 | SSI | 379 | 1.16 (0.95 –1.43) | 0.15 |
| IL-8 | AMI | 376 | 1.04 (0.87 –1.25) | 0.70 |
|  | Delirium | 376 | 1 (0.88 –1.14) | 0.96 |
| IL13 | AKI | 383 | 0.88 (0.72 –1.07) | 0.20 |
| Lymphocytes | AKI | 566 | 1.01 (0.98 –1.04) | 0.60 |
| MCP-1 | AKI | 389 | 0.99 (0.87 –1.12) | 0.87 |
|  | AMI | 389 | 0.95 (0.84 –1.07) | 0.44 |
|  | Delirium | 389 | 0.96 (0.88 –1.05) | 0.39 |
| Neutrophils | AKI | 566 | 1 (0.97 –1.03) | 0.88 |
|  | AMI | 566 | 0.99 (0.96 –1.02) | 0.57 |
| sIL-2Ra | AF | 381 | 1.02 (0.93 –1.12) | 0.71 |
|  | Delirium | 381 | 1.04 (0.91 –1.19) | 0.58 |
| TLR2 | AKI | 2438 | 1 (0.96 –1.04) | 0.87 |
|  | Delirium | 2438 | 0.99 (0.97 –1.01) | 0.37 |
| TLR4 | AKI | 2438 | 0.99 (0.95 –1.03) | 0.46 |
|  | Delirium | 2438 | 1.01 (0.99 –1.03) | 0.41 |
| TNF-a | AF | 379 | 1.04 (0.94 –1.15) | 0.39 |
|  | AMI | 379 | 1.02 (0.85 –1.23) | 0.82 |
|  | Delirium | 379 | 1.01 (0.88 –1.16) | 0.94 |
|  | SSI | 379 | 1.14 (0.93 –1.4) | 0.21 |
